# Complications and Status Upgrades among Adult Heart Transplant Candidates with Durable LVADs: Waiting 6 to 8 Years for Status Escalation Is Too Long

**DOI:** 10.1101/2025.09.22.25336215

**Authors:** Daniel J Ahn, Antony Attia, Toshihiro Nakayama, Nikhil Narang, Kiran K Khush, William Parker, Kazunari Sasaki

## Abstract

**Introduction:** After the 2018 allocation policy change, the rate of listings and transplants with durable LVADs has decreased significantly in favor of bridging patients from temporary mechanical circulatory support to heart transplant. The Organ Procurement and Transplantation Network (OPTN) recently approved a policy, to be implemented in September 2026, stipulating that patients supported by durable LVADs for 6 and 8 years will obtain statuses 3 and 2, respectively.

**Methods:** Using OPTN data, we identified all adult heart transplant candidates with a durable LVAD implanted between October 18, 2018 and May 31, 2025. We estimated the cumulative incidence of status upgrades and durable LVAD-related complications, treating transplantation and waitlist removal before experiencing complications as competing events. We also assessed how the composition of the adult heart transplant waitlist on June 1, 2025 would have changed based on the upcoming policy change.

**Results:** During the study period, 3,881 adult patients were listed for heart transplant with a durable LVAD. 3,182 (82.0%) of the durable LVADs were Abbott HeartMate 3, 568 (14.6%) were Medtronic Heartware HVAD, and 91 (2.3%) were Abbott HeartMate II. Transplant centers submitted a total of 6,924 justifications for status upgrades due to LVAD-related complications (6.3% status 1, 34.3% status 2, and 59.4% status 3) for 1,500 (38.6%) of these patients, with a median of 3 per patient. The cumulative incidence of complications or status upgrades was 38.6% [95% CI (37.1%, 40.2%)]. Nearly all of the 2,381 patients who did not experience any complication or status upgrade during listing were removed from the waitlist by 6 years. Had the upcoming OPTN policy change been implemented on June 1, 2025, the proportion of the waitlist that would have achieved higher priority status instantaneously was 0.06%.

**Conclusions:** The cumulative incidence of status upgrades and complications among heart transplant candidates with durable LVADs was nearly 40% within 6 years of device implantation. The upcoming OPTN policy to escalate patients to statuses 3 and 2 after 6 and 8 years of durable LVAD support, respectively, is unlikely to make a meaningful impact on waitlist priority status.

## Introduction

Since October 2018, the United States donor heart allocation system has rank-ordered adult transplant candidates using six ordinal statuses.^1^While patients who are clinically stable and supported by durable left ventricular assist devices (LVADs) used to have the second-highest waitlist priority before October 2018, they are now listed as status 4 as a default. As a result, rates of durable LVAD implantation, especially as a bridge to transplantation, have decreased significantly,^2–4^ coupled with a significant increase in listings with temporary mechanical circulatory support (tMCS) devices such as intra-aortic balloon pumps (IABPs), percutaneous endovascular ventricular assist devices (PEVADs), and extracorporeal membrane oxygenation (ECMO).^5,6^

In response, the Organ Procurement and Transplantation Network (OPTN) recently approved a policy supporting prioritization of durable LVAD candidates to statuses 3 and 2 after 6 and 8 years of durable LVAD support, respectively.^7^ While the concept of a “time-served” exception to afford durable LVAD patients a greater chance at obtaining a transplant was well-received, many patients, advocacy groups, and transplant programs voiced concerns that waiting for 6 and 8 years is too long, with candidates at high risk of experiencing device-related complications during this time.^7^ In this analysis, we aimed to determine the rate at which patients with durable LVADs obtain status upgrades or experience complications as well as the expected impact of the upcoming policy change on the distribution of patients on the waitlist.

## Methods

### Data Source and Study Population

This study used data from the Organ Procurement and Transplantation Network (OPTN) Standard Transplant Analysis and Research (STAR) files. The STAR files include data on all donors, waitlisted patients, and transplant recipients in the United States. The Health Resources and Services Administration (HRSA) provides oversight to the activities of the OPTN contractors. This study was exempted by the Stanford University Institutional Review Board. We identified all adult heart transplant candidates on the waitlist who received a durable LVAD between October 18, 2018 and May 31, 2025, with follow-up available until June 30, 2025.

### Patient Characteristics

We collected relevant demographic and clinical data from patients at listing. Variables included sex, age at durable LVAD implantation, body mass index (BMI), race, blood type, primary diagnosis, insurance type, and functional status as approximated by the Karnofsky performance scale. We categorized Karnofsky performance scale into good (80%-100%), moderate (50%-70%), and poor (10%-40%), as was done previously.^8^ In addition, we categorized BMI into the following: underweight (<18.5 kg/m^2^), normal (18.5-24.9 kg/m^2^), overweight (25-29.9 kg/m^2^), and obese (≥30 kg/m^2^).

### Outcomes and Statistical Analysis

The primary outcomes of our study were rates of heart transplantation and complications/status upgrades related to durable LVAD support. As described in OPTN policy, specific LVAD-related complications such as device malfunctions, device infections, and mucosal bleeding meet standard listing criteria.^1^ However, similar to prior research,^9^ we considered any upgrade in status above status 4 among patients with durable LVADs to be due to a complication. For example, we reasoned that even though IABP and PEVAD support (meeting status 2 criteria) or VA-ECMO support (meeting status 1 criteria) are not explicitly categorized as complications, patients who are originally clinically stable with durable LVADs at status 4 but then require tMCS or any other type of support meeting elevated status criteria should be considered as having failure or complication of the durable LVAD. The only status upgrade criterion that we did not consider to be a complication was the 30 days of discretionary status 3 time for durable LVAD patients. This specific categorization of status 3 is not due to failure or complication of the device. It is rather intended to be a mechanism for providing elevated priority for a limited time while these patients remain clinically stable.^1^

We then estimated the cumulative incidence of experiencing a complication/status upgrade by 6 years after durable LVAD implantation. We treated heart transplantation and waitlist removal before experiencing status upgrades/complications as competing events. We stratified our analysis by type of durable LVAD placed and performed Fine-Gray analysis for all competing events. Patients were censored if their durable LVAD was explanted before transplant. We also identified all patients who received a transplant with a durable LVAD still in place and plotted the distribution of their statuses at transplant. We compared the median time from durable LVAD implantation to transplant between patients who did or did not experience any complications/status upgrades with Mood’s median test. We then obtained all status justifications submitted on behalf of patients who obtained a status higher than 4 at any time during listing after durable LVAD implantation.

Finally, we looked at the composition of the adult heart transplant waitlist on June 1, 2025 and determined how the distribution of candidates, by status, would have changed instantaneously under three different policy changes. The first policy change is phase 1 of the upcoming September 2026 OPTN policy in which candidates who have had a durable LVAD for 6 and 8 years would receive status 3 and 2, respectively. The second policy change is phase 2 of the aforementioned policy, in which candidates with an LVAD for 5 and 7 years would get statuses 3 and 2, respectively. In reality, phase 2 of the policy would be implemented in early 2028.^7^ The final policy change is a hypothetical scenario in which candidates with a durable LVAD for 2 and 4 years would receive status 3 and 2, respectively. We chose 2 years for status 3 due to recent evidence demonstrating that post-transplant survival decreases significantly among patients transplanted with a HeartMate 3 after 2 years of durable LVAD support.^10^ We then chose 4 years for status 3 simply by adding 2 years, as was done for both phases 1 and 2 of the upcoming policy. This specific analysis includes all patients with durable LVADs on the waitlist on June 1, 2025, including those with devices that were implanted before October 18, 2018.

To compare demographic and clinical data of patients stratified by type of durable LVAD, we performed descriptive statistics with chi-square tests for categorical variables and Wilcoxon rank-sum tests for continuous variables. All statistical tests were two-sided, and we considered a p-value of < 0.05 to be significant. We performed all analyses with R (version 4.5.1). Complete statistical code necessary to reproduce the study results is available online.^11^

## Results

A total of 3,881 adult heart transplant candidates had a durable LVAD placed between October 18, 2018 and May 31, 2025. 3,182 (82.0%) of the durable LVADs were Abbott HeartMate 3 and 570 (14.7%) were Medtronic Heartware HVAD. A full list of the durable LVAD types is provided in **Supplemental Table 1. Table 1** summarizes the demographic and clinical characteristics of all patients stratified by type of durable LVAD. Patients with the HeartMate 3 durable LVAD were more likely to be older at listing (56 vs 54 years, p < 0.001) and to be male (78.2% vs 72.7%, p = 0.002). There were no significant differences in race, BMI, blood type, primary diagnosis, and insurance type. Non-ischemic dilated cardiomyopathy was the most common diagnosis overall (61.9%).

**Table 1:**
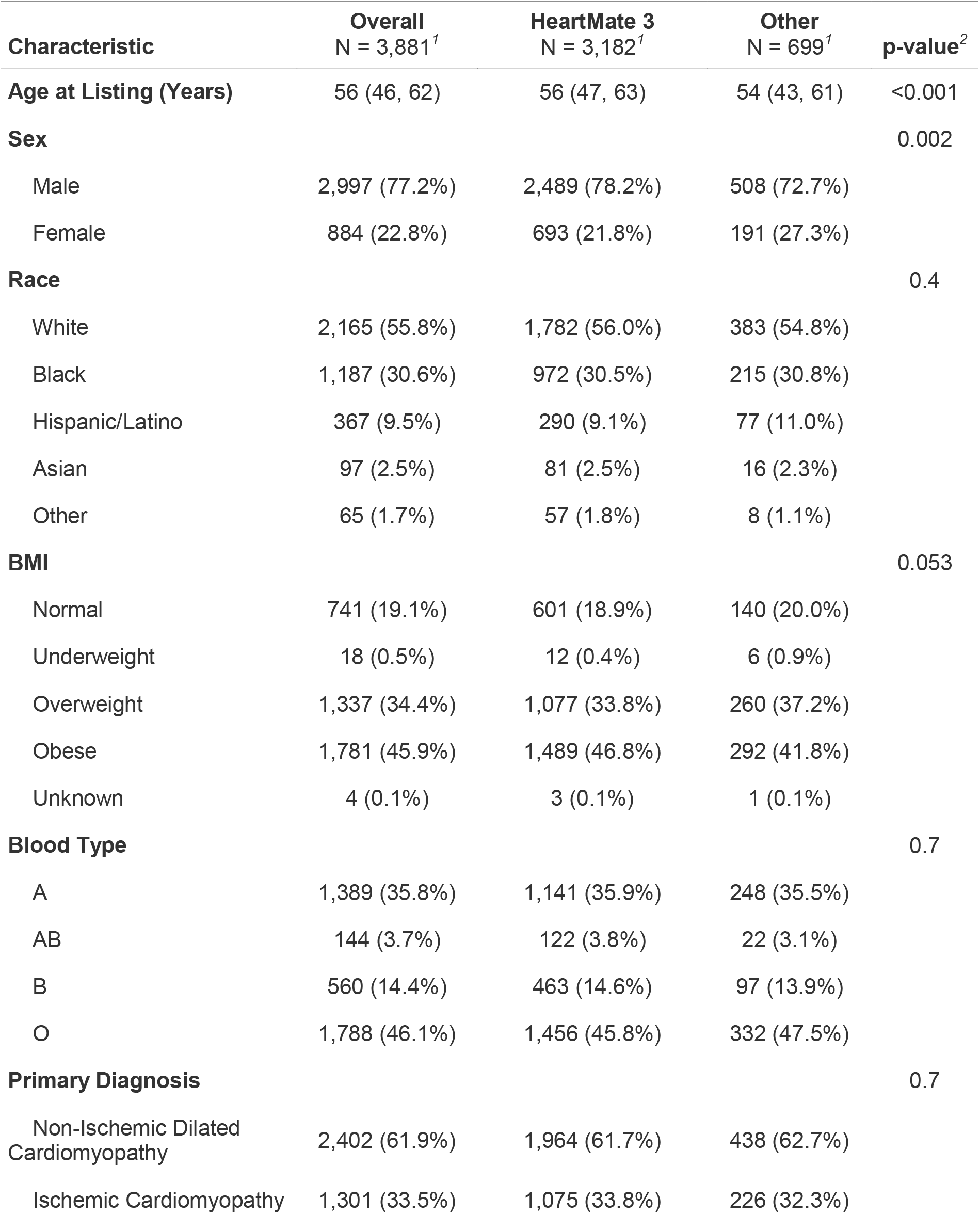
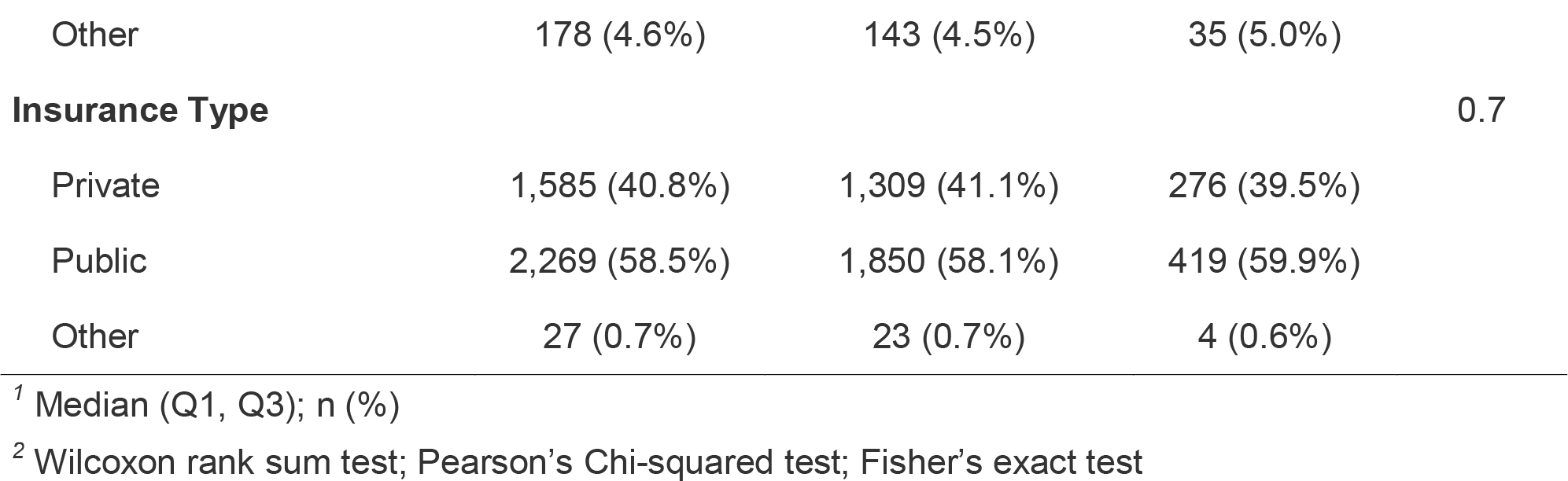
Demographic and Clinical Characteristics of Study Cohort, Stratified by Durable LVAD Type.

### Complications and Status Upgrades

Transplant centers submitted a total of 6,924 justification files for status upgrades/LVAD-related complications for 1,500 patients (38.6%), with a median of 3 file submissions per patient. Reasons for status upgrades are listed in **Table 2**, stratified by type of durable LVAD. Overall, the most common reasons for status 1 upgrades were exceptions (52.8%) and placement of a non-dischargeable surgical BIVAD (20.5%). For status 2, the most common reasons were exceptions (65.1%) and device malfunction (14.5%). The majority of status 3 upgrade requests were due to device infections (58.6%). The types of complications and status upgrades differed significantly by durable LVAD type (all p < 0.001). 6,907 (99.8%) of the justification files were submitted within 6 years of durable LVAD implantation, and the median time to first submission of a file for status upgrade was 440 days (IQR 182–847 days).

**Table 2:** Durable LVAD Complications or Status Upgrades During Listing, Stratified by Device Type.

| Characteristic | Overall<br>N = 6,924<sup>1</sup> | HeartMate 3<br>N = 5,197<sup>1</sup> | HVAD<br>N = 1,383<sup>1</sup> | Other Durable<br>LVAD<br>N = 344<sup>1</sup> | p-value<sup>2</sup> |
| --- | --- | --- | --- | --- | --- |
| **Reason for Status 1 Upgrade** | N = 439 | N = 361 | N = 64 | N = 14 | <0.001 |
| Device Recall | 16 (3.6%) | 0 (0.0%) | 16 (25.0%) | 0 (0.0%) |  |
| ECMO | 18 (4.1%) | 14 (3.9%) | 3 (4.7%) | 1 (7.1%) |  |
| Exception | 232 (52.8%) | 189 (52.4%) | 35 (54.7%) | 8 (57.1%) |  |
| MCS with Life Threatening Ventricular Arrhythmia | 83 (18.9%) | 74 (20.5%) | 4 (6.3%) | 5 (35.7%) |  |
| Non-Dischargeable Surgical BIVAD | 90 (20.5%) | 84 (23.3%) | 6 (9.4%) | 0 (0.0%) |  |
| **Reason for Status 2 Upgrade** | N = 2,373 | N = 1,847 | N = 409 | N = 117 | <0.001 |
| Device Malfunction | 345 (14.5%) | 229 (12.4%) | 82 (20.0%) | 34 (29.1%) |  |
| Device Recall | 167 (7.0%) | 14 (0.8%) | 141 (34.5%) | 12 (10.3%) |  |
| Exception | 1,544 (65.1%) | 1,350 (73.1%) | 150 (36.7%) | 44 (37.6%) |  |
| IABP | 11 (0.5%) | 7 (0.4%) | 3 (0.7%) | 1 (0.9%) |  |
| Non-Dischargeable LVAD | 7 (0.3%) | 5 (0.3%) | 1 (0.2%) | 1 (0.9%) |  |
| PEVAD | 38 (1.6%) | 15 (0.8%) | 2 (0.5%) | 21 (17.9%) |  |
| Recurrent or Sustained VT/VF | 32 (1.3%) | 30 (1.6%) | 1 (0.2%) | 1 (0.9%) |  |
| TAH, RVAD, VAD for Single Ventricle Patients, or BiVAD | 229 (9.7%) | 197 (10.7%) | 29 (7.1%) | 3 (2.6%) |  |
| **Reason for Status 3 Upgrade** | N = 4,112 | N = 2,989 | N = 910 | N = 213 | <0.001 |
| Continuous IV Inotropes | 13 (0.3%) | 11 (0.4%) | 2 (0.2%) | 0 (0.0%) |  |
| Device Infection | 2,408 (58.6%) | 2,091 (70.0%) | 225 (24.7%) | 92 (43.2%) |  |

| **Characteristic** | **Overall**<br>**N = 6,924<sup>1</sup>** | **HeartMate 3**<br>**N = 5,197<sup>1</sup>** | **HVAD**<br>**N = 1,383<sup>1</sup>** | **Other Durable**<br>**LVAD**<br>**N = 344<sup>1</sup>** | **p-**<br>**value<sup>2</sup>** |
| --- | --- | --- | --- | --- | --- |
| Device Recall | 415 (10.1%) | 7 (0.2%) | 388 (42.6%) | 20 (9.4%) |  |
| Exception | 525 (12.8%) | 382 (12.8%) | 73 (8.0%) | 70 (32.9%) |  |
| MCS with Aortic Insufficiency | 128 (3.1%) | 121 (4.0%) | 5 (0.5%) | 2 (0.9%) |  |
| MCS with Hemolysis | 29 (0.7%) | 0 (0.0%) | 16 (1.8%) | 13 (6.1%) |  |
| MCS with Mucosal Bleeding | 58 (1.4%) | 47 (1.6%) | 8 (0.9%) | 3 (1.4%) |  |
| MCS with Pump Thrombosis | 363 (8.8%) | 161 (5.4%) | 189 (20.8%) | 13 (6.1%) |  |
| MCS with Right Heart Failure | 173 (4.2%) | 169 (5.7%) | 4 (0.4%) | 0 (0.0%) |  |

<sup>1</sup> n (%)
<sup>2</sup> Fisher's Exact Test for Count Data with simulated p-value (based on 2000 replicates)
Abbreviations:
LVAD (Left ventricular assist device)
ECMO (Extracorporeal membrane oxygenation)
MCS (Mechanical circulatory support)
BIVAD (Biventricular assist device)
IABP (Intra-aortic balloon pump)
PEVAD (Percutaneous endovascular ventricular assist device)
TAH (Total artificial heart)
RVAD (Right ventricular assist device)
VAD (Ventricular assist device)

The cumulative incidence of experiencing any complication or status upgrade (other than status 3 for discretionary LVAD time) at 6 years was 38.6% [95% CI (37.1%, 40.2%)], which was higher than that of transplantation at status 4 or status 3 for discretionary LVAD time [cumulative incidence 37.1%, 95% CI (35.6%, 38.6%)] (**Figure 1**). Patients with HeartMate 3 were significantly more likely to obtain a transplant (p = 0.002) and significantly less likely to experience a complication or status upgrade (p < 0.001) than those with non-HeartMate 3 durable LVADs (**Figure 2**). Out of 2,381 candidates who did not experience any complications, only 4 had not been removed from the waitlist by 6 years after durable LVAD placement.

**Figure 1:**
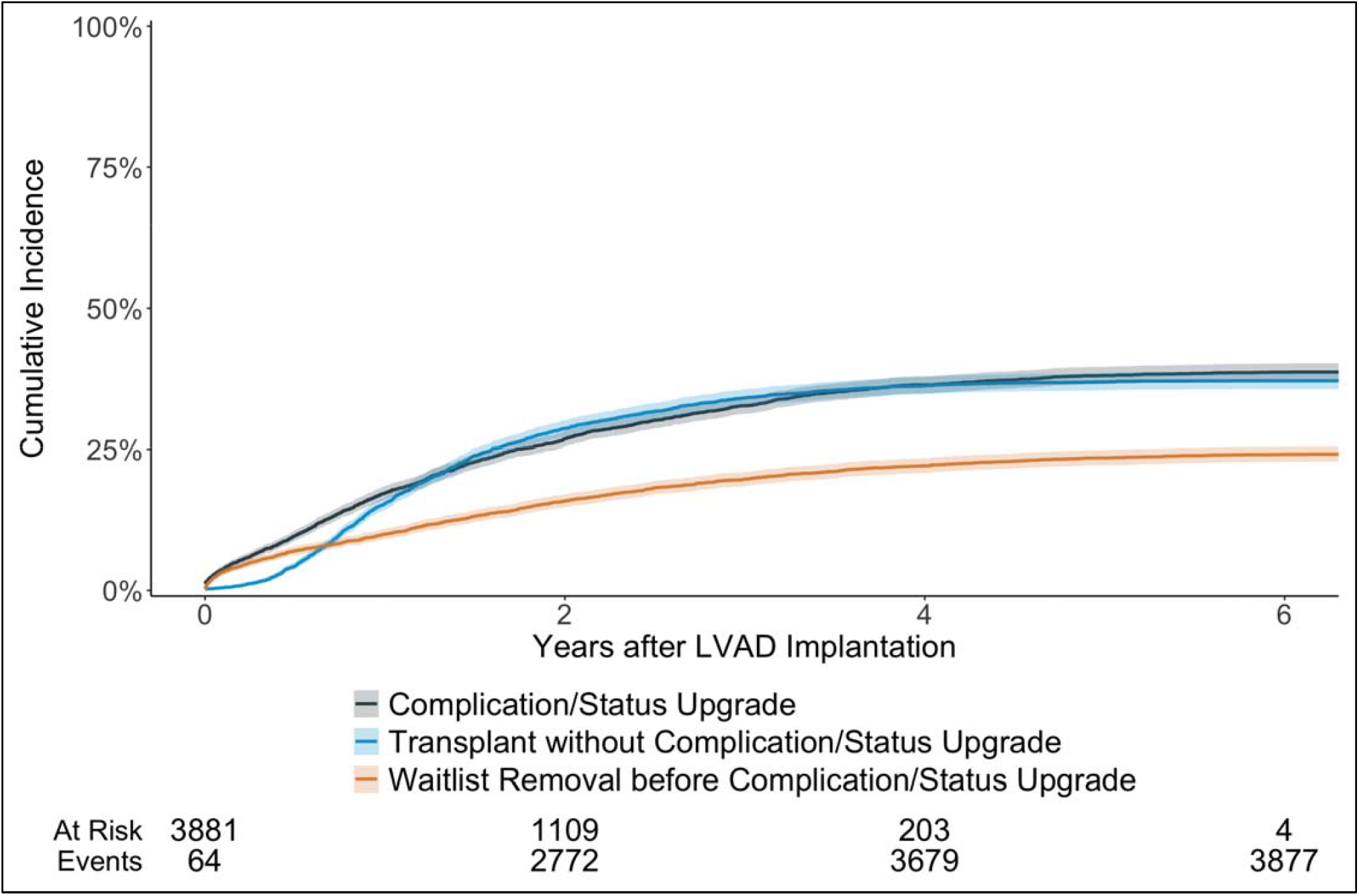
Cumulative incidence of complication or status upgrade (other than status 3 for discretionary LVAD time), treating transplant and waitlist removal (other than transplant) prior to complication/status upgrade as competing events

**Figure 2:**
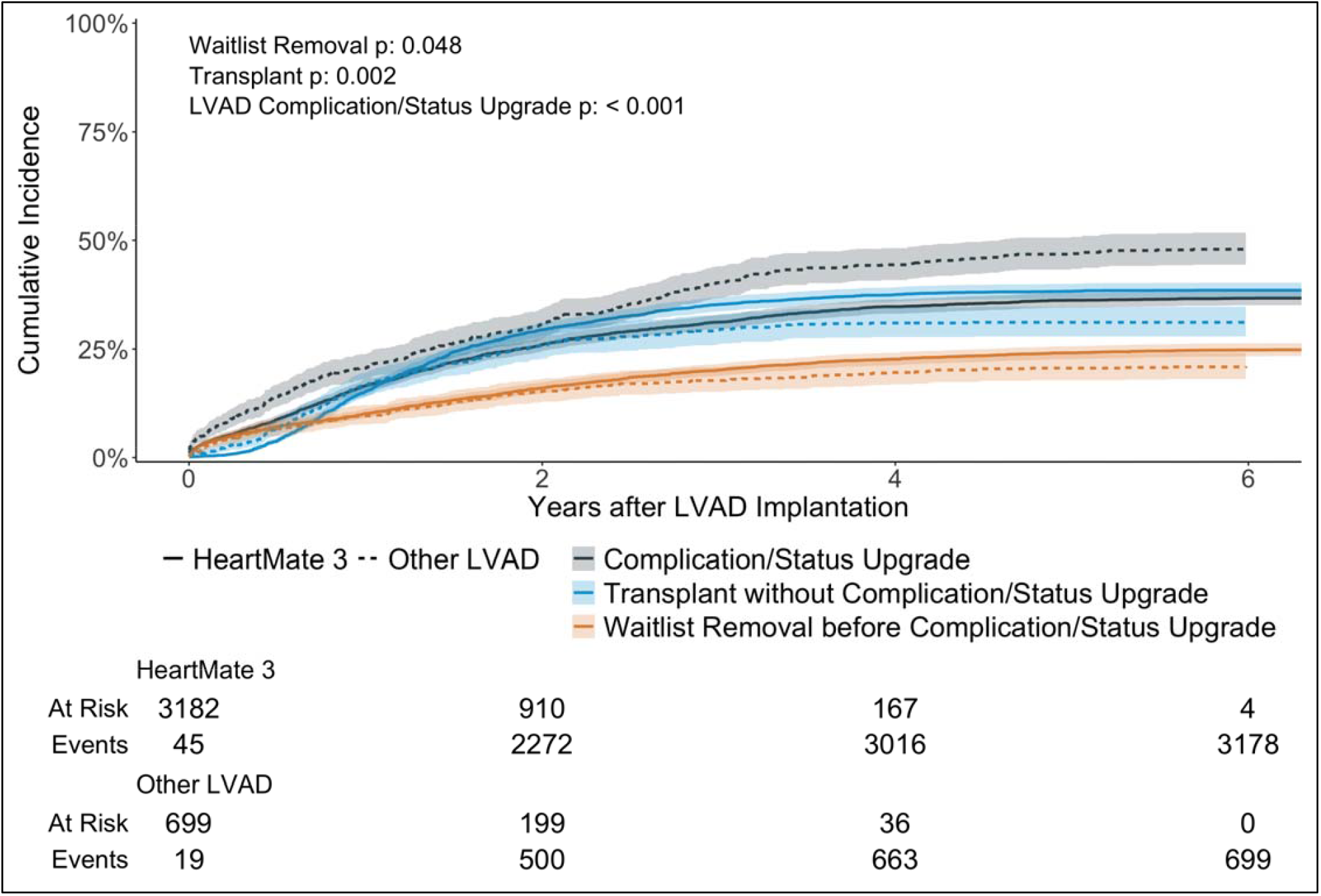
Cumulative incidence of complication or status upgrade (other than status 3 for discretionary LVAD time) stratified by durable LVAD type. This treats transplant and waitlist removal (other than transplant) prior to complication/status upgrade as competing events. Fine-Gray results for all competing events are in the top left corner of the figure.

### Heart Transplants with Durable LVADs

2,703 (69.6%) patients received a heart transplant with a durable LVAD in place, and the distribution of statuses they had at transplant are available in **Figure 3**. 218 (8.1%) were status 1, 642 (23.8%) were status 2, 982 (36.3%) were status 3, and 861 (31.8%) were status 4. 579 (59.0%) of the patients with status 3 were exercising their 30 days of discretionary LVAD time. The median time to transplant for candidates who only had status 4 or status 3 for discretionary LVAD time during listing was 429 days (IQR 267–694 days) and that for patients who had experienced a complication or status upgrade at least once while waitlisted was 549.5 days (IQR 234.25–985.25 days) (p < 0.001).

**Figure 3:**
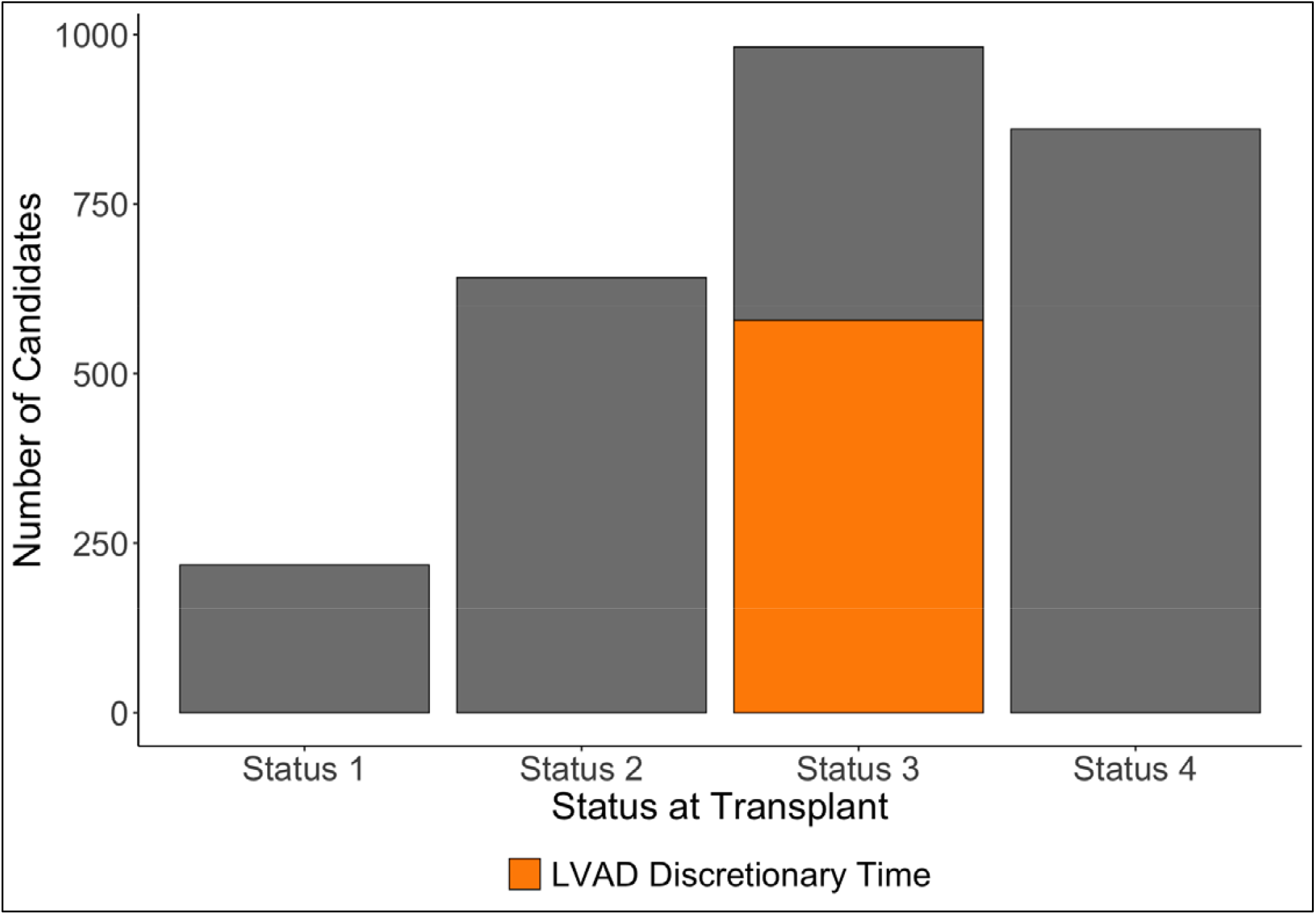
Distribution of status levels of 2,703 patients with durable LVADs at the time of transplantation. Status 3 is stratified by whether patients met status 3 criteria with discretionary LVAD time.

### Composition of the Adult Heart Waitlist

On June 1, 2025, there were 3,214 patients on the adult heart waitlist, of whom only 80 (2.5%) had a durable LVAD (**Figure 4**). Of the 80 durable LVADs, 72 (90%) were HeartMate 3, 4 (5%) were Heartware HVAD, and 4 (5%) were HeartMate II. 12 (15.0%) of the 80 patients were inactive. 34 (50.0%) of the 68 active patients were already listed at statuses 1 to 3. If phase 1 of the upcoming September 2026 policy change had been implemented on June 1, 2025, there would have been no additions to status 3, and only 2 patients with status 4 would have been added to status 2 instantaneously. If phase 2 of the policy had been implemented, an additional 3 patients would have been added to status 3. If a hypothetical policy in which patients with a durable LVAD for 2 and 4 years would obtain statuses 3 and 2, respectively, were implemented on June 1, 2025, 5 patients with status 4 would become status 3, and 16 patients with status 4 would become status 2 instantaneously. When examining the composition of the overall waitlist (**Figure 5**), the proportion of the listed candidates that would have changed statuses with phase 1 of the accepted policy would be 2 out of 3,214 (0.06%), phase 2 of the accepted policy would be 5 out of 3,214 (0.16%), and the hypothetical 2- and 4-year policy would be 21 out of 3,214 (0.65%).

**Figure 4:**
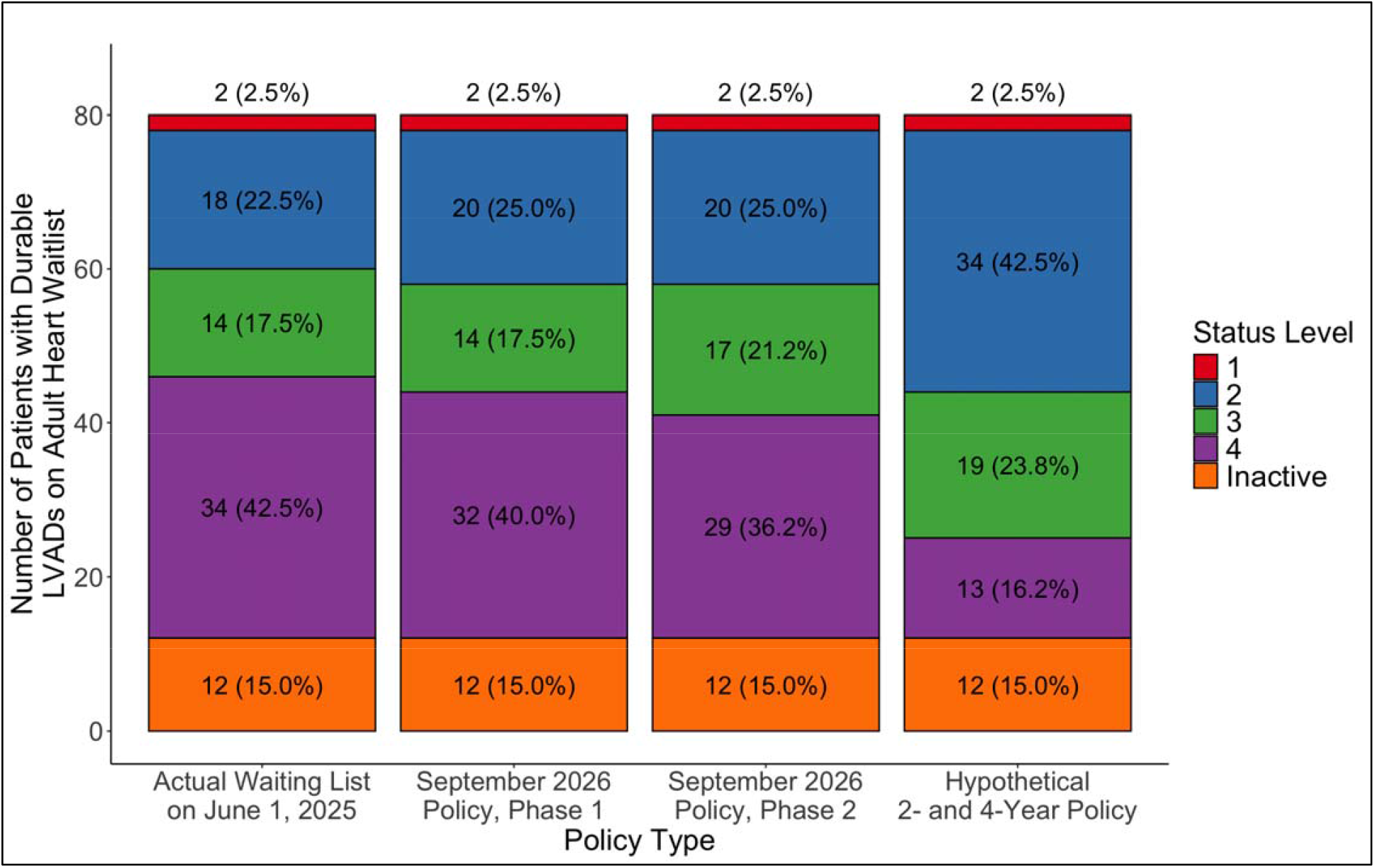
Distribution of all patients with durable LVADs on the waitlist on June 1, 2025. The first column is the actual distribution. The second and third columns show the distributions with phases 1 and 2 of the upcoming OPTN policy in September 2026. The last column depicts how the distribution would change instantaneously if patients obtained statuses 3 and 2 after 2 and 4 years with a durable LVAD, respectively.

**Figure 5:**
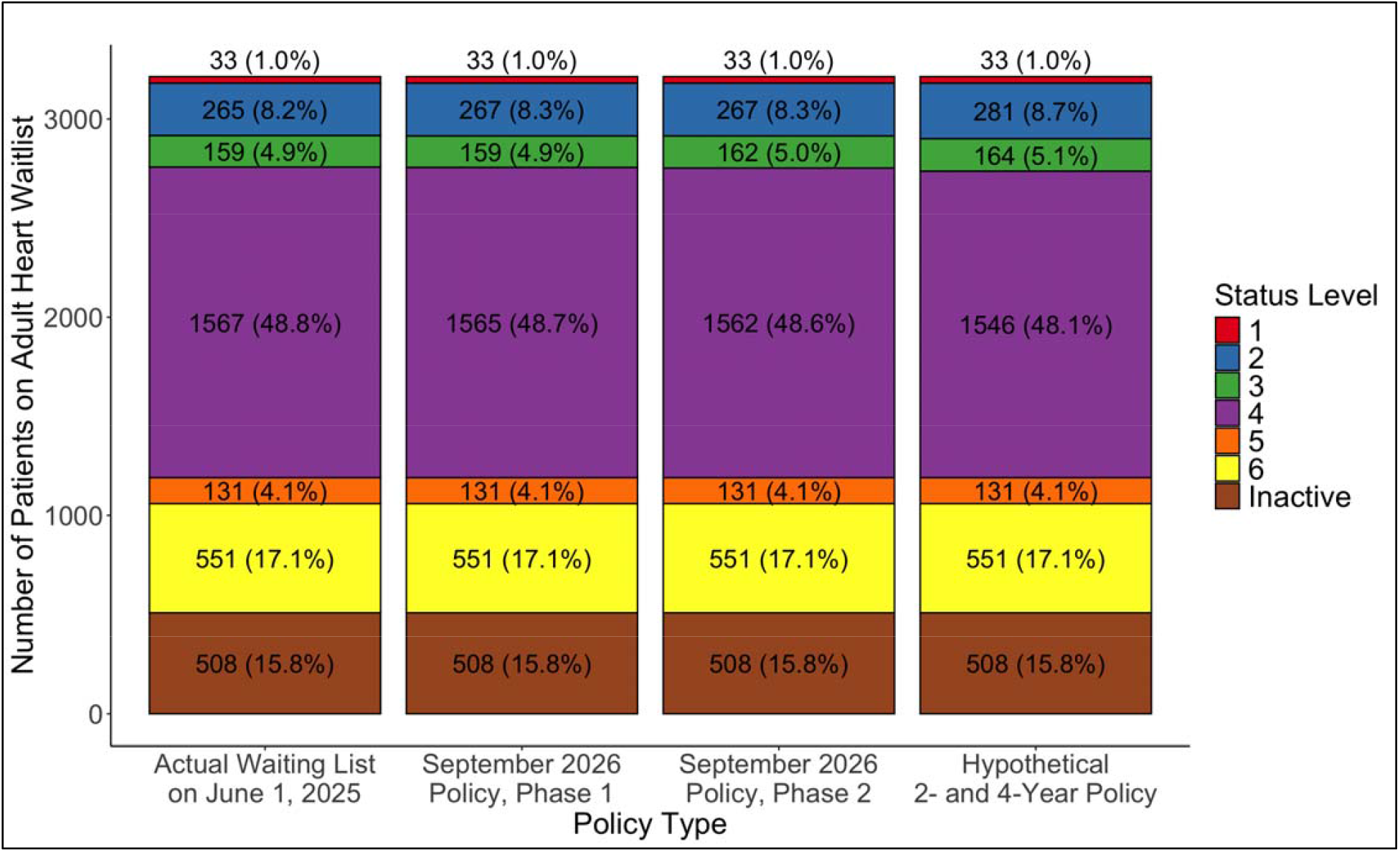
Distribution of the whole adult heart transplant waitlist on June 1, 2025. Same as in Figure 4, the first column is the actual distribution. The second and third columns show the distributions with phases 1 and 2 of the upcoming OPTN policy in September 2026. The last column depicts how the distribution would change instantaneously if patients obtained statuses 3 and 2 after 2 and 4 years with a durable LVAD, respectively.

## Discussion

In this registry-based study of adult heart transplant candidates supported by durable LVADs awaiting heart transplantation in the US, we report the following findings: 1) the cumulative incidence of complications or status upgrades was nearly 40% by 6 years after LVAD implantation, 2) nearly 100% of all complications or status upgrades that occurred in the study cohort happened by 6 years, and 3) the composition of the adult heart waitlist in early June 2025 would not have changed significantly had the upcoming OPTN policy change been implemented then.

Despite advances in durable LVAD technology that have conferred lower morbidity and mortality,^12–15^ the number of patients listed with durable LVADs has decreased significantly.^4,16^ Only if patients develop device-related complications, such as refractory infections and pump malfunctions, can they be upgraded from status 4 to a higher status.^1^ As a result, transplant centers have moved away from durable LVADs while simultaneously increasing use of tMCS as a bridge-to-transplant strategy, despite current OPTN policy recommending centers to transition patients from tMCS devices to durable LVADs unless there are absolute contraindications.^1^ In particular, rates of IABP utilization rose more than 3-fold after the allocation policy change in 2018.^17^ While the OPTN has recognized the dwindling numbers of patients with durable LVADs awaiting heart transplant and will implement a “time-served” policy in September 2026, our results show that 6 and 8 years are too long to wait for a status upgrade.

The OPTN Heart Committee wrote this policy based on an analysis of the adult heart waitlist as of April 30, 2024 and aimed to avoid a large influx of high priority listings that may negatively impact the waitlist survival of other candidates already listed as status 2 and 3.^7,18^ Thus, the committee decided on 6 and 8 years because these timeframes limited the total number of candidates with durable LVADs switching instantaneously to statuses 3 and 2, compared to other potential proposals.^7^ However, April 30, 2024 was more than a year ago, and in our analysis of the waitlist as of June 1, 2025, only 68 out of more than 3,000 patients had a durable LVAD with an active status, with half already at a waitlist priority higher than status 4. We show that if phase 1 of the accepted policy had been implemented on June 1, 2025, only 2 candidates would have switched statuses instantaneously. Even with a policy proposing 2 and 4 years of waiting, the total proportion of candidates who would have changed statuses is less than 1%. A simulation or retrospective match run analysis is needed to estimate the true impact of any such policy on the waitlist mortality of other candidates. However, given the steadily decreasing number of candidates with durable LVADs since October 2018^4^ and the fact that less than 3% of waitlisted patients in early June 2025 had durable LVADs, we predict that by the time the policy is implemented in September 2026, not only will there be very few candidates with durable LVADs left on the waitlist but the estimated impact of the policy will be miniscule.

Furthermore, we argue that a timeline of 6 and 8 years is too long not only because of low predicted policy impact but also because many patients experience adverse events leading to higher statuses and other outcomes long before 6 years. Nearly 100% of all status upgrades/complications submitted by transplant centers in our study cohort occurred within 6 years of durable LVAD implantation. Additionally, the policy is intended to benefit candidates who are stable with a durable LVAD,^7^ but 2,377 out of 2,381 patients who never experienced complications or status upgrades during the study period had already been removed from the waitlist by 6 years after durable LVAD implantation. Moreover, recent findings have shown that post-transplant survival among patients with durable LVADs is significantly worse when they are transplanted with complications meeting criteria for status 3 and 2 compared to when they are clinically stable with status 4.^9^ In the same vein, post-transplant survival may be worse following 2 years of durable LVAD support among patients with HeartMate 3 devices.^10^ Both the amount of time spent with durable LVAD support and complications/status upgrades experienced during support from durable LVADs may have significant impact on patients’ waitlist survival, morbidity, and even post-transplant survival; our results provide compelling evidence that waiting 6 and 8 years is too long.

The OPTN Heart Committee had attempted to create a policy based on when the pre-transplant medical urgency of candidates with durable LVADs would match that of patients with statuses 3 and 2.^7^ However, we argue that applying such a framework for comparing medical urgency may not be appropriate when determining the priority of durable LVAD patients because they are typically stable and have low risk of mortality when they are free from complications. Instead, one solution to determine when patients should be prioritized above status 4 is by estimating the duration of device support after which the survival benefit afforded by both a durable LVAD and transplant is maximal.

### Limitations

There are several potential limitations to our study. First, we defined complications and status upgrades based off the status justification files submitted by transplant centers. Furthermore, we did not include temporary inactivations as complications despite evidence that inactivations are typically related to poor functional or clinical status,^19^ which could be indicative of complications related to durable LVAD support. As a result, we may have underestimated how many complications actually occurred in this population.

Second, we defined all upgrades above status 4 except status 3 for discretionary LVAD time as complications even if they are not explicitly categorized as LVAD-related complications in OPTN policy. However, any adverse clinical event that causes patients with durable LVADs to meet criteria for increased waitlist priority above what is afforded by status 4 should be considered as a complication. Exceptions accounted for a significant portion of the status upgrades within this cohort. While they are associated with lower medical urgency in general,^20,21^ OPTN policy^1^ states that patients with approved exceptions in statuses 1 to 3 must be admitted to the hospitals where they are listed. We argue that inpatient admission with a durable LVAD should be considered an adverse event.

Third, we only included patients who had durable LVADs placed after October 2018 in our cohort, which may have limited our study population. However, we restricted our cohort because we wanted to study the status upgrade and complication rate of recently placed durable LVADs, particularly the HeartMate 3, in order to maintain clinical relevance. Furthermore, by limiting our study population to durable LVAD placement after October 2018, we may have had limited follow-up. However, very few patients in our cohort were censored with nearly everyone experiencing either a complication/status upgrade or removal from the waitlist either for transplant or another reason.

### Conclusions

The cumulative incidence of status upgrades and complications among heart transplant candidates with durable LVADs listed as status 4 was nearly 40% by 6 years after implantation. The upcoming policy change in September 2026 to escalate patients to statuses 3 and 2 after 6 and 8 years of durable LVAD support, respectively, is unlikely to have significant impact.

## Data Availability

All data utilized for analysis in the present study are available upon reasonable request to the Organ Procurement and Transplantation Network. The code written to perform all analyses in this manuscript are available here at this GitHub repository: https://github.com/danieljaechulahn/determining-timing-of-lvad-priority

## Abbreviations

tMCS: Temporary mechanical circulatory support
IABP: Intra-aortic balloon pump
PEVAD: Percutaneous endovascular ventricular assist device
LVAD: Left ventricular assist device
HRSA: Health Resources and Services Administration
OPTN: Organ Procurement and Transplantation Network

## Author Contributions

Dr. Ahn and Dr. Sasaki had full access to the data in the study and take responsibility for the integrity of the data and the accuracy of the data analysis.

*Concept and design:* Ahn, Khush, Parker, Sasaki

*Acquisition, analysis, or interpretation of data:* Ahn, Nakayama, Attia, Sasaki

*Drafting of the manuscript:* Ahn

*Critical review of the manuscript for important intellectual content:* All authors.

*Statistical analysis:* Ahn

*Obtained funding:* Parker

*Administrative, technical, or material support:* Parker, Sasaki.

*Supervision:* Khush, Parker, Sasaki

## Disclaimer

The interpretation and reporting of these data are the responsibility of the authors and in no way should be seen as an official policy of or interpretation by the OPTN or the US government.

