## Supplemental Table 1 for "Complications and Status Upgrades among Adult Heart Transplant Candidates with Durable LVADs: Waiting 6 to 8 Years for Status Escalation Is Too Long"

| **Characteristic** | **N = 3,881***^1^* |
| --- | --- |
| Durable LVAD Brand |  |
| HeartMate 3 | 3,182 (82.0%) |
| HeartMate II | 92 (2.4%) |
| Heartware HVAD | 570 (14.7%) |
| Evaheart | 6 (0.2%) |
| ReliantHeartAssist 5 | 2 (0.1%) |
| Heartsaver VAD | 4 (0.1%) |
| Worldheart Levacor | 3 (0.1%) |
| Unknown (Categorized as  Durable LVAD) | 22 (0.6%) |
| *^1^* n (%) | |
